# Clinical trial emulation can identify new opportunities to enhance the regulation of drug safety in pregnancy

**DOI:** 10.1101/2021.11.12.21266269

**Authors:** Anup P. Challa, Xinnan Niu, Etoi A. Garrison, Sara L. Van Driest, Lisa M. Bastarache, Ethan S. Lippmann, Robert R. Lavieri, Jeffery A. Goldstein, David M. Aronoff

## Abstract

From the perspective of most regulatory agencies, it is usually unethical to perform interventional clinical trials on pregnant people. While this policy recognizes the vulnerability of an expectant mother and unborn child, it has created a public health emergency for millions of pregnant patients through a dearth of robust safety data for many common drugs. To address this problem, we harnessed an enterprise collection of 2.8M electronic health records (EHRs) originally collected from routine primary care, leveraging the data linkage between mothers and their babies to create a surrogate for randomized, controlled drug trials in this population. To demonstrate the feasibility of our clinical trial emulation platform to stimulate new hypotheses for post-market drug surveillance, we identified 1,054 drugs historically prescribed to pregnant patients and developed a medication history-wide association study and follow-up evidence synthesis platform—leveraging expert clinician review and real-world data analysis—to test the effects of maternal exposure to these drugs on the incidence of neurodevelopmental defects in their children. Our results replicate known teratogenic risks and existing knowledge on drug structure-related teratogenic risks. Herein, we highlight 5 common drug classes that we believe warrant further assessment of their safety in pregnancy. We also discuss our efforts to develop a discovery-to-regulatory framework that could allow for pragmatic translation of our results to enhanced regulatory policy. Collectively, our work presents a simple approach to evaluating the utility of EHRs in guiding new regulatory review programs focused on improving the delicate equipoise of accuracy and ethics inherent to assessing drug safety in an extremely vulnerable patient population.

## Introduction

At the point of care, pregnant patients are a vulnerable population: physicians must exercise caution in prescribing many common drugs to these patients, given the risks of toxicity for their developing fetuses^1^. However, consideration of fetal toxicity in drug development is largely nonsystematic. While teratogenicity scores established by regulatory agencies like United States Food and Drug Administration (FDA) are discrete, these criteria provide little concrete distinction among score classes, making it difficult for drug developers to accurately gauge the fetal toxicity risks of a molecule^2^. FDA’s updated teratology assessment guidelines in the 2014 Pregnancy and Lactation Labeling Rule aimed to increase the contextual relevance of developmental toxicity evaluation, but this guidance has been slow to translate to evaluative change at the point of care, which remains largely aligned with the previous five-pronged letter scale^3,4^. The result is a vicious cycle that promotes the approval of drugs without adequate data on their safety and efficacy in pregnant populations, as expectant patients are routinely and ethically excluded from clinical trials, out of concern for fetal harm upon exposure to drugs with uncertain, pre-clinical teratogenicity data. In fact, of 213 new drugs approved by FDA between 2003 and 2012, only 5% contained human data in the pregnancy section of their labels^5^. Herein, pregnant patients may not receive benefit from available medications, given obstetricians’ cautious fears of causing harm to their patients’ fetuses^6^. These factors have created a substantial gap in knowledge on pharmacotherapy for diseases during pregnancy, which has resulted in undertreatment of chronic and acute illnesses in pregnant people, while also increasing potential risk of harm to their fetuses.

Therefore, the inability to generate new drug safety and efficacy information in pregnant patients through prospective experiments like randomized, controlled trials (RCTs) underscores the urgent need for new methods to ethically assess this information, to improve quality of care for these underserved patients, and to ensure health equity for this vulnerable population through enhanced drug product labelling and marketing. Such an opportunity for discovery of new drug safety insights for pregnant patients may be available through strategic analysis of large numbers of existing healthcare documents like electronic health records (EHRs) that were collected during routine patient care. Collectively, EHRs can uniquely replicate the natural history of pregnancy by linking medical information of pregnant patients and their neonates, such as mothers’ prescriptions (while expectant) and the perinatal diseases of their children^7–9^. This information allows for the creation of a unique framework of relational knowledge generation. Namely, a surrogate for RCTs in pregnant patients can result from stratifying EHR data into distinct cohorts by patients’ documented exposure—or lack thereof—to a drug of interest and subsequently developing an inferential model to relate incidences of maternal drug exposure and neonatal disease^8^. While these experiments are not a replacement for prospective clinical studies, such a platform of clinical trial emulation presents an ethical way of studying the effects of drug exposure in pregnant people with human data, on a significant scale and across all drug classes.

Existing literature that describes the safety of most drugs potentially prescriptible in pregnancy remains overwhelmed by conflicting studies—the majority of which only present results from pre-clinical animal models of drug testing and the minority of which are empirical case reports or case series among relatively few patients^10^. Deciding to prescribe a drug to a pregnant patient involves balanced evaluation of the patient’s need for treatment (drug efficacy) and the risk of injury to the patient’s fetus (drug safety). However, providers cannot make these informed decisions without robust and definitive safety data.

Previous work that has attempted to clarify knowledge on drug safety in pregnant patients has relied on observational and retrospective analyses of databases like public insurance claims, measuring the significance in coincidence of a neonatal disease of interest and prescription of a drug of interest to the neonates’ mothers^7,11^. While these studies have added new—and often valuable—narratives of drug safety to the literature, our research is innovative because it uses EHR data, attempts relational inference, and probes such drug-disease relationships at scale. Collectively, these factors allow us to advance the ontological reliability and epistemological robustness of data-driven studies of adverse pregnancy outcomes^8^.

Our research makes novel use of a database of 2.8M EHRs at Vanderbilt University Medical Center (VUMC) to curate our trial emulation cohorts. The data innovation in studying EHR data over evaluating public insurance claims is that this choice mitigates significant demographic biases (e.g., poverty) that are present within public payor records. Overcoming the effects of such potentially confounding variables requires the integration of advanced methods of propensity scoring (PS) to properly evaluate the coincidence of maternal drug exposure and pediatric disease, which defies the key algorithmic design principle of parsimony and results in poor model performance^12^. In contrast, VUMC is an urban medical center that features a demographically diverse patient population, as previous studies using these EHR data affirm^13^. Indeed, self-reporting patient registries—another popular choice for observational data to study health outcomes in pregnancy—are also inherently limited in their integrity, as patients are often unreliable historians of their own care^14^. In contrast, our study promotes data integrity by studying provider-maintained healthcare information.

Technical innovation in this project also rests within the rigor of the analytical methods we employ (Figure 1, below)^8^. We apply a mode of systematic, relational inference to maternal drug exposure and perinatal disease that we believe is more directly and appropriately aligned with the etiology of drug-associated birth defects, compared to the highly coincidental frameworks that dominate the literature. We achieved inference suggestive of causality through harmonizing the phenome-wide association (PheWAS), which was originally developed at VUMC to discover genetic links to clinical phenotypes, with a rigorous, standardized consensus prioritization approach that considered clinical practice and RCT data to move from data-based associations towards etiology discovery^15^. By developing a medication history-wide association study (MedWAS) to suggest pharmacological determinants of neonatal diseases, we optimized on algorithms that underlie PheWAS to explore nascent patterns across the drug-disease hypotheses that our model revealed. In this way, we used MedWAS as a novel method of clinical trial emulation^8^. Target trials are an epidemiological method of retrospective data analysis that make use of existing clinical information and high-powered statistical algorithms to create artificial subject profiles from all relevant and available patient data within a cohort of interest. This curation then allows for relational analysis of subjects’ drug histories against a morbidity of interest, facilitating potential simulation of a clinical trial when prospective experiments are not feasible^16–18^. The approach in this manuscript alludes to a target trial by following similar approaches to data curation and stratification, statistical inference, and outcomes prioritization, though unlike the archetypal target trial developed by Hernán and Robbins for claims data and consortial data banks^16^, our distributed workflow for practicably applying trial emulation to a single health system’s mother-baby EHRs means that some aspects of our procedure rely on manual evidence synthesis, rather than harnessing end-to-end automation. To our knowledge, there have been very few (and relatively small) RCT emulation projects evaluating pregnant patients^19^, allowing us to innovate in exploring the power of this novel method at scale^8,20^.

**Figure 1:**
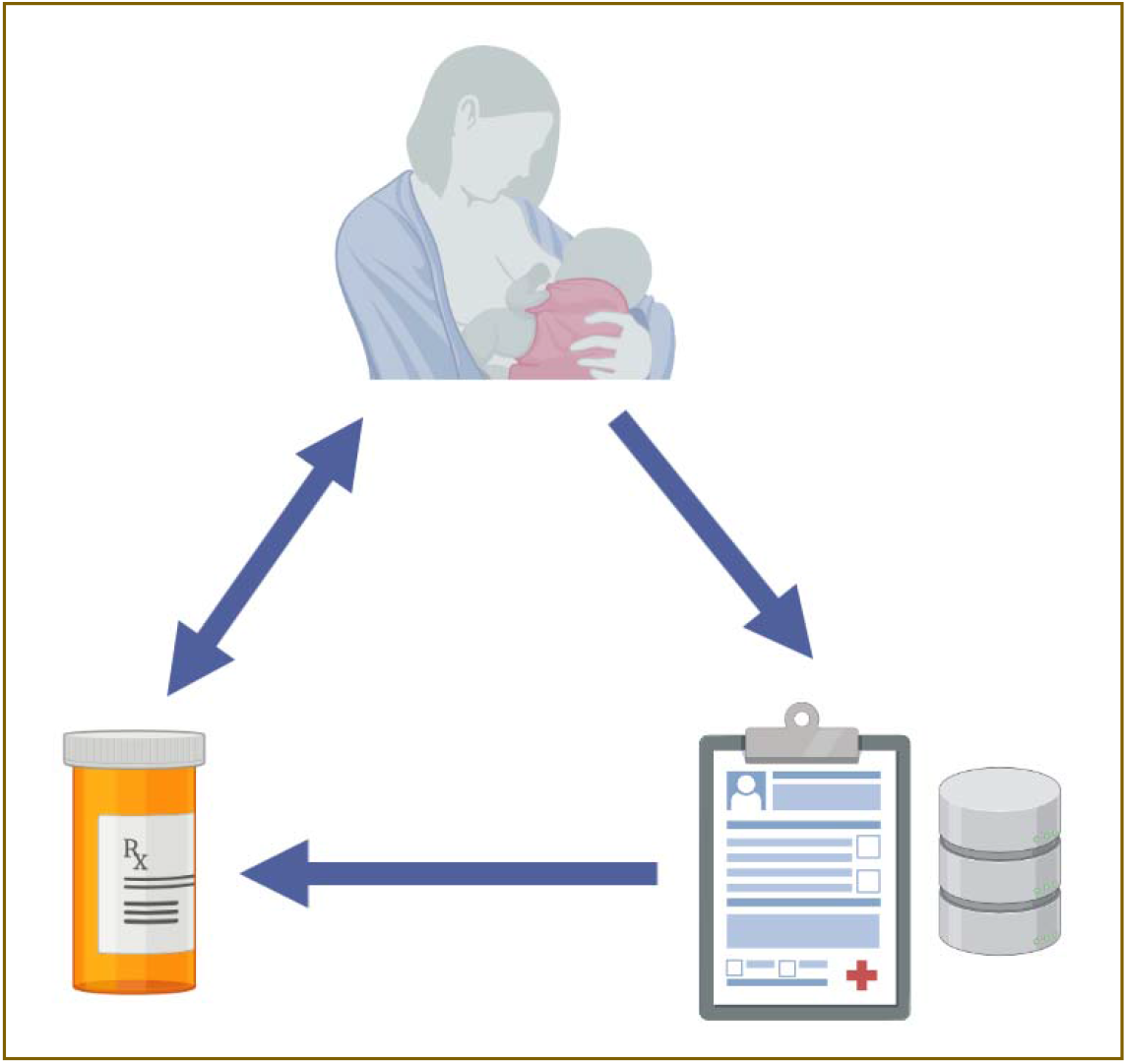
As a powerful framework of retrospective analysis, clinical trial emulation could allow pregnant people and their children to benefit from drug safety insights that arise from analysis of their own medical records.

Using MedWAS, we present systematic RCT simulation across all drugs prescribed to pregnant people and all diseases within neonatal EHRs at VUMC: herein lies the conceptual innovation of our approach. Historically, researchers studying the safety of pharmacotherapy in pregnancy with statistical methods have communicated through a “one drug—one disease—one publication” model. While this practice provides bandwidth for deep interrogation of a single drug-disease hypothesis, it further diversifies the pool of existing data that remains conflicting and inconsistent, since the methods in such papers can become overfitted for studying the safety of other drugs that are prescriptible in pregnancy. In contrast, our approach is sufficiently powered to analyze maternal prescriptions and neonatal diseases across a large healthcare enterprise. In this way, our platform provides a reproducible framework of drug safety assessment that may be sufficiently generalizable to accommodate testing of new drugs. We are unaware of such a drug-agnostic and phenotype-agnostic model in the available literature on drug safety in pregnancy.

We have a record of work in using statistical methods like PheWAS to generate strong hypotheses of efficacy for new drug development^9,21,22^. Here, we apply that expertise to construct MedWAS as an innovatively scalable, RCT-emulatory approach for the surveillance of drug safety in pregnancy. We also present potential avenues for complementarity between MedWAS and our previous attempts to develop a machine learning (ML) approach capable of identifying chemical structures that predispose drugs towards increased teratogenic risk when prescribed during pregnancy^23^.

## Methods

The approach that we describe below is an explanatory summary of the data preprocessing (for cohort selection) and informatics procedures (for drug-disease testing) that we provide in cookbook format within the “Supplementary Information” that accompanies this manuscript. A diversity and inclusion report for the maternal and neonatal EHRs we analyzed is also included in the “Supplementary Information” accompanying this manuscript.

We tested the hypothesis that an RCT emulation platform can effectively establish relational inference between mothers’ exposures to drugs with uncertain safety and perinatal diseases in their neonates. In establishing the feasibility of our tool to accomplish post-market drug surveillance, we restricted ourselves to the analysis of only neurological morbidities as a base case, given that the ontologies that codify these diseases have strong bases of relational logic^24^. We expect the general framework of the analytical and signal evaluation procedures we present here will be analogously applicable to interrogation of neonatal diseases in other organ systems.

The Institutional Review Board (IRB) of Vanderbilt University approved the research that we describe below as exempt from human subjects research (IRB #191553).

### Cohort Selection and Data Curation

To mimic the enrollment of pregnant patients in a drug safety trial, we used ML to curate and block appropriate treatment and control (drug-exposed vs. not drug-exposed) cohorts across all 1,054 agents that are documented as prescriptions to pregnant patients in eStar, VUMC’s EHR system. A listing of these agents is available in “Supplementary Information.” To select our cohorts, we probed VUMC’s Research Derivative (RD), a database of fully identified clinical and administrative information from 2.8M patients that contains data like International Classification of Disease-9/10 (ICD-9/10) billing codes (which codify nearly all existing human morbidities), patient demographics, lab results, medications, and clinical narratives from five different relational health information systems that source directly from patient care^25,26^. To effectively create trial emulation cohorts across the agents we probed from these data, we first established the following phenotyping rule as inclusion criteria for patient “enrollment” in treatment and control groups: Population: RD; Include: Mom/baby link (1 or more), where specified medication (1 or more where date during mother EHR pregnancy=yes) and clinic note in baby EHR suggests record of care (1 or more postpartum).

Herein, our criteria for allocating pregnant patients to a drug treatment group required baseline, confirmed pregnancy among all candidate mothers, with a record of at least one prescription of the specified drug in the mother’s EHR during their gestational period and successful delivery of a neonate who received their own EHR at VUMC (such that their health outcomes were available for our analysis). Defining pregnancy and gestational period in a systematic way from the EHR remains a non-standardized analytical practice and therefore required us to develop an inferential approach reliant on a data dictionary of relevant ICD-10 codes for gestational period. For interested readers, we describe this approach in “Supplemental Information.” We designed our inclusion criteria to maximize the data available to our model, so we could achieve the highest power for demonstrating preliminary proof-of-concept for our approach. Herein, we harnessed downstream evidence synthesis to vet our trial outcomes, rather than establishing very tight inclusion (and exclusion) criteria *a priori* to mitigate confounding variables.

Next, we leveraged a suite of natural language processing (NLP) tools to extract phenotypic attributes and maternal drug exposures from narrative EHR data among all patients within the 94,872 EHRs (48,434 mother-baby EHR pairs) that met our inclusion criteria for at least one study drug. These tools included a general-purpose NLP tool (KnowledgeMap concept identifier (KMCI)^27,28^), ML-based clinical note section tagger (SecTag^29,30^), and MedEx, an NLP algorithm for identifying medication exposures within free clinical text^29,31^. KMCI identifies Unified Medical Language System concepts^32^ using a shallow parser, word sense disambiguation, and semantic regularization, and includes a module to identify negation^27^. MedEx uses context-free grammar and a rule-based approach to extract detailed medication information (including dose, frequency, and route) from free text. MedEx encodes an ingredient barcode for all drugs, such that drug mentions extracted from EHRs are continuously linked to existing drug ontologies from which additional pharmacological data may be mined (e.g., RxNorm concept unique identifier^33^)^29,31^. These standardized systems have been used to process more than 60 million documents at Vanderbilt and elsewhere. Here, we used them to capture all drug mentions and available ICD-9/10 codes and to facilitate requisite matching of free-text disease terms to concept unique identifiers for candidate mothers and their linked neonates, as well as to extract all available demographic information for “enrolled” mothers and babies. Enacted across all combinations of diseases and maternal drug histories in our population, our workflow enabled the curation and stratification of patient data to empower >1.7M combinatorial RCT simulations, as we describe below.

### Implementation of MedWAS

PheWAS is a common, systematic ML approach to identify novel associations between disease and genetic variants and to discover pleiotropy using EHR data linked to DNA. It is a method that scans phenomic data for genetic associations using Phecodes mapped to ICD-9/10 codes from the EHR. Multiple publications demonstrate that PheWAS is a feasible method to rapidly generate novel hypotheses on the underpinnings of disease^15,34–37^. We repurposed the PheWAS framework to develop an innovative MedWAS, in identifying the extent to which the perinatal phenotypes in our cohorts are plausibly related to exposure to the drugs in each simulated trial’s treatment group. Herein, our proof-of-concept MedWAS model took an input of babies’ neurological diseases from all mother-baby cohorts we constructed and outputted the maternal medication exposures putatively related to babies’ phenotypes. While it is easiest to envision our trial platform through the canonical stratification of mother-baby cohorts by maternal drug exposure, our adoption of neonatal disease-contingent inference across treatment-defined maternal cohorts allowed us to develop capacity for discovery of multiple drug exposures as etiologies for our phenotypes of interest.

MedWAS operated in direct analogy to PheWAS by using its component logistic classification methods (logit) to identify neonatal disease as a function of maternal exposure to a drug of interest and by reporting a *p*-value for each of these drug-disease tests that reflected the strength of logit alignment after correction for multiple testing of a drug across all neonatal diseases in our cohorts. In doing this across 1,054 native maternal drug exposures and the neurological subset of 1,678 EHR-embedded phenotypes—first, on a pilot-scale, with 5.7K EHR pairs, and subsequently on our full data set of 49K mother-baby dyads—each trial was controlled by cases of neonatal disease linked to pregnant patients without a record of exposure to the test drug. Herein, we also computed an odds ratio (OR) as a proxy for the effect size of hypothetical drug-disease enrichment across each of our tested case and control populations. Because there are known associations among the representations of input and output data and PheWAS model performance^35–37^, we iteratively assessed MedWAS performance with several standard representations of the drug and disease data (i.e., different levels of Anatomical Therapeutic Chemical (ATC) codes for drug entities^38^ and Phecodes and ICD-9/10 codes for diseases^39^) from our cohorts to prevent confounding of our results by data type. Figure 2, below, summarizes the premise of our MedWAS approach. The list of 1,678 Phecodes we employed is publicly accessible through the open-source code for the PheWAS package (see https://github.com/PheWAS/PheWAS).

**Figure 2:**
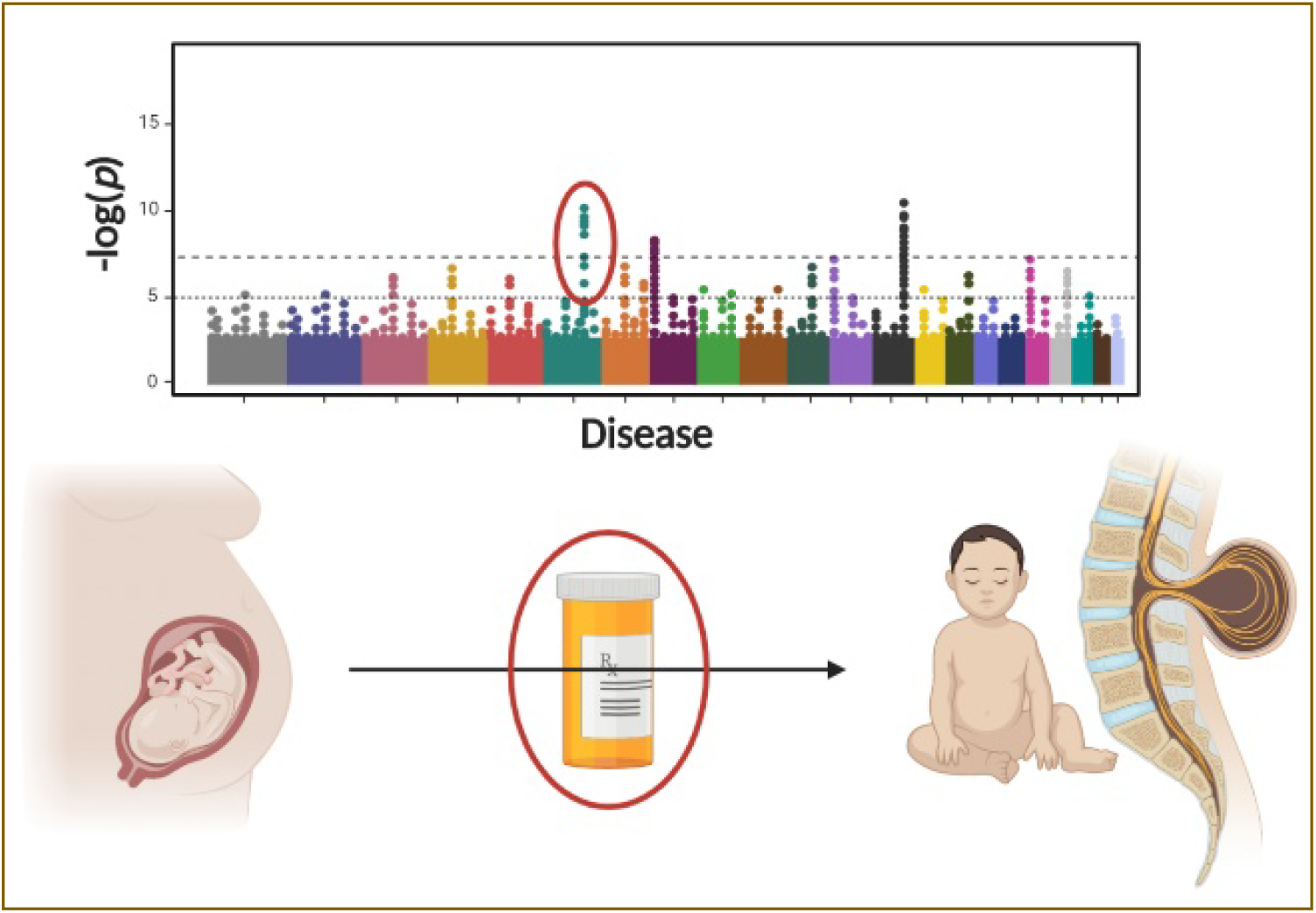
MedWAS allowed us to ethically discover new drug safety insights from clinical trial-emulatory analysis of mothers’ drug histories and their neonates’ diseases.

### Evidence Synthesis and Hypothesis Prioritization

While the explicit goal of our work was to establish a platform for generating hypotheses of drug safety that may be pursued in more targeted studies in the future, we affirm that a non-deterministic challenge in pursuing our experiments was accurate prioritization of MedWAS’s predicted drug-disease relationships by their clinical, biological, and statistical plausibility, given the number of RCT simulations we executed rapidly within our analytical framework. We attempted to meet this challenge by ranking our results with the following heuristics: concordance with known fetal safety risks from published drug labels, a soft constraint of Bonferroni significance (with correction from baseline *p* ≤ 0.05) and OR > 1, compelling clinical reviews from obstetrician and pediatrician consults on the plausibility of substantially implicated drug prescriptions and teratogenic outcomes, reproducibility between MedWAS outputs and the results from our previous work that identified drug structures linked to adverse birth outcomes^23^, and evidence against “confounding by indication” from harmonizing systematic chart review of mothers’ baseline disease states with knowledge of known vertical disease transmission risks within our treatment cohorts. Our application of the *p*-value as a soft prioritization constraint that complemented systematic review from our clinical stakeholders aligns with guidance to this effect from American Statistical Association^40^.

To parse MedWAS results we believed were not clinically plausible or were potentially confounded, we began by restricting all signals associated to nutraceutical products, as we recognized that patient history-informed capture of food and nutritional supplement use data in the EHR is highly unreliable. These agents are available over-the-counter (OTC) and often incompletely reported by patients, such that mention of the agent does not always imply true exposure during gestation^41^. Then, we consulted a pediatrician with expertise in clinical pharmacology on our study team to identify neurological Phecodes with unlikely manifestation in the perinatal period; these diseases were mainly neurocognitive (e.g., dyslexia) and therefore excluded from consideration as true model results. Our pediatrics consult further stratified higher-level versions of the phenotype embeddings in our model outcomes as incident in infants, toddlers, school-age children, or adolescents, based on disease pattern presentations from clinical practice. Consequently, we excluded all outcomes not plausibly detectable in infants.

Following our pediatrician’s review, we consulted a practicing obstetrician on our study team, who has training in clinical pharmacology and maternal-fetal medicine, to identify the plausibility of prescription of the drugs implicated in our model during pregnancy. In completing this review, our obstetrics consultant synthesized knowledge from her own prescriptive practice, prescriptive guidelines from American College of Obstetricians and Gynecologists, Society for Maternal-Fetal Medicine, departmental practice guidelines at Vanderbilt, and clinical decision software (CDS) like UpToDate^4^ and Reprotox^42^ to stratify our signals as “high-yield” and “low-yield” outcomes. We defined high-yield outcomes as those which demonstrated statistical significance, at least 1% co-incidence rate between drug prescription and pediatric disease (such that, with our sample sizes of mothers prescribed each drug and neonates born with each disease, we prioritized only non-unary outcomes), and unclear prescriptive recommendations and/or practice guidelines for implicated drugs (e.g., FDA score C and conflicting case reports described in CDS). These drugs also had plausible prescription during the first trimester of pregnancy, when the majority of neurological organ development occurs. Low-yield outcomes included signals rooted in drugs available OTC, such that EHR data on drug use were not reliable for our first-pass analysis, and signals with drugs sparsely prescribed to pregnant patients in the United States of America due to lack of regional drug supply and/or existing guidance against prescription of these drugs during pregnancy. Our consideration of the latter revealed to us that our low-yield signals may be artifactual noise from our inferential approach to defining gestational period, if these drugs appeared in pregnant patients’ EHRs before discontinuation, when providers first learned of their patients’ pregnancies.

Our designation of the yields of our signals were powered by a spreadsheet model we developed, which codified the considerations above by fields including the following. In an *ad hoc* fashion, both consultants, as well as a pharmacologist, removed drugs from consideration which presented with implausible pharmacokinetics for their associated toxicities (e.g., non-systematic absorption).

- “drug’s original indication” (to help identify potential cases of confounding by maternal morbidity)
- “FDA drug class”
- “trimester of prescription”
- “intrapartum or immediate postpartum prescription?” (a response of “yes” to this question resulted in a signal’s relative de-prioritization, given our interest in antepartum exposures and the difficulty of perfectly ascertaining gestational period within the EHR)
- “duration of prescription”

Figure 3, below, provides a summary of our process for developing and vetting MedWAS data.

**Figure 3:**
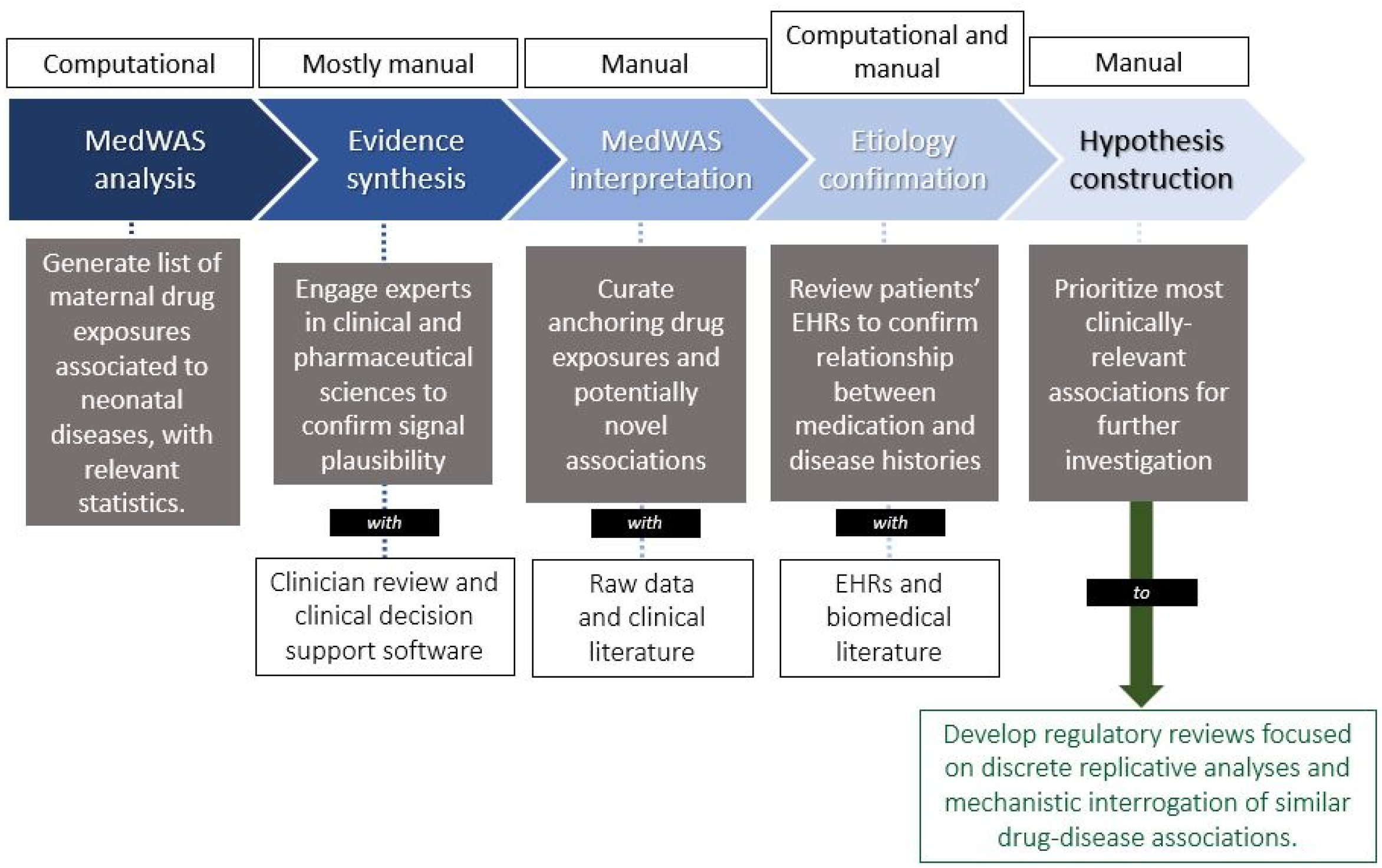
Summary of the process to develop and assure the quality of MedWAS signals—the goal of our approach is to unearth and prioritize drug safety risks, towards generation of the highest quality hypotheses to inform new regulatory review programs. Engagement of obstetric, pharmacological, and regulatory stakeholders is inherent to this process.

## Data Availability Statement

Disclosure of our MedWAS data, though de-identified and aggregated, is subject to approval and oversight by the Office of Contracts Management (OCM) at VUMC, as our source data is derived from protected health information (PHI), and some drug-disease pairs are individually identifiable. Therefore, institutional policies prevent us from publicly releasing our data tables and their annotations, but, within the data sharing regulations of our institution, we have attempted to provide meaningful information on the content and formatting of our outputs throughout this manuscript. We are committed to open-source science and to ensuring the reproducibility of the research we present here; therefore, we are happy to discuss data transfer requests with researchers interested in our results. Interested investigators should contact the Corresponding Authors at the addresses accompanying this manuscript, and they are happy to discuss forwarding such requests to OCM.

## Results and Discussion

We present MedWAS as a simple and flexible process of generating novel hypotheses for post-market drug surveillance of drug safety in pregnancy, which takes strategic advantage of the milieu of primary medical care for pregnant patients and the data routinely generated through these encounters. The approach also relies on known patterns of prescriptive behavior in pregnancy to emulate—at limited capacity—baseline cohort randomization before testing the effects of drug exposure. While we cannot claim “full” randomization, we acknowledge the following assumptions towards a level of randomized assignment of drug vs. no drug for pregnant patients with disease: first, prescriptive strategies for these patients are likely heterogeneous, given limited safety data and providers’ differing risk-benefit evaluations of prescriptions. Therefore, not all diseased patients will receive pharmacotherapy, resulting in inherent randomization. We acknowledge that even with this assumption, an exact, 50%-50% split in drug treatment is unlikely to manifest. Second, undue bias towards treatment at baseline is unlikely to exist (i.e., providers are not naturally “instructed” to treat all ailing pregnant patients with drugs), so the lack of a prior treatment probability supports the randomization described above. Coupled with the controlling inherent to each MedWAS test, this randomization allows us to claim foundational RCT emulation via MedWAS; we present key results from our platform below, along with a discussion of the advantages, several limitations, and positive reception of our attempt, which we believe collectively define new opportunities for expansion of our approach as a systematic attempt at drug safety assurance that is powered by real-world evidence (RWE).

### Proof of Concept

*Prima facie*, we consider MedWAS successful for its robust capacity to accommodate the largescale testing that we envisioned: following our experimental design, pilot testing, and localized sensitivity analyses, we executed 1,770,290 drug-disease experiments using a high-performance cluster with 2,400 processor cores hosted by the Southern Crossroads server^43^ for supercomputing.

As we describe in “Methods,” facing an abundance of generated data, we restricted analysis of the reliability of our results to a single physiological system, to allow for deep contextual analysis. Accordingly, we selected to analyze 1,414 neuroteratogenic signals meeting our aforementioned definition of statistical significance, given expertise in neuropathogenesis within our study team and the spatially and temporally focal nature of many neurodevelopmental anomalies to neurulation^44^, which occurs in the first trimester of pregnancy^45^. In analyzing this functional area, we assume that our insights are sufficiently generalizable to similar physiology in other organ systems, but we also acknowledge that signals among systemic developmental phenotypes may require more formal network analyses. In considering the validation procedures we describe in “Methods” and the evidence requirements we discuss below for signal confirmation, we found that MedWAS performed best on the bases of ATC-4 and Phecode representations of our drug and disease data, respectively. Choosing these representations allowed us to balance data granularity and utility in optimizing model performance, as we tested associations of agent names (but not formulations, as would be available from ATC-5 embeddings) against high-level phenotype codes with logical mappings to the ICD ontology. While drug formulation could present interesting relationships to toxicity (e.g., through elevated concentrations at sensitive physiological sites like the cervix), we consider that our inability to capture this information does not detract from the power of our model to robustly capture associations between maternal drug exposures and adverse neonatal outcomes, as the explicit goal of our model was to discover relationships between the agents mothers consume and adverse outcomes in their neonates. In this way, we consider formulation to have trace effects on fetal toxicity, further given that most agents within our list of agents are consumed orally.

We observed replication of 8 well-known teratogens [phenytoin^46^, valproate^47^, fenofibrate^48^, quinapril^49^, retinoids (tazarotene, vitamin A, and adapalene)^50^, and, topiramate^51^] and 2 teratogens confirmed by our clinical consults [salicylates (phenyl salicylate and salicylic acid)^52,53^] within our MedWAS results. We consider the according 22 signals (presented, in part, within Table 1, below) as positive population controls for our model: when we identified maternal medication history across our health system, we anticipated that such “anchoring” drugs would present with associations to neuroteratogenic outcomes. Negative population controls (i.e., prescription drugs with known protective effects against teratogenicity and/or zero baseline risk of teratogenic outcomes) are inherently uncommon and were therefore difficult for us to develop, further given that protective agents like folate are often taken by all expectant mothers receiving medical care during pregnancy, in addition to other potentially toxic drugs^54^. Herein, our replication of positive control signals through MedWAS allowed us sufficient confidence to procced with analysis of our model outcomes; our intention to develop structured statistical models with inherent controlling—both for each drug-disease test and across our population—also affirms our non-exploratory study design.

**Table 1*:**
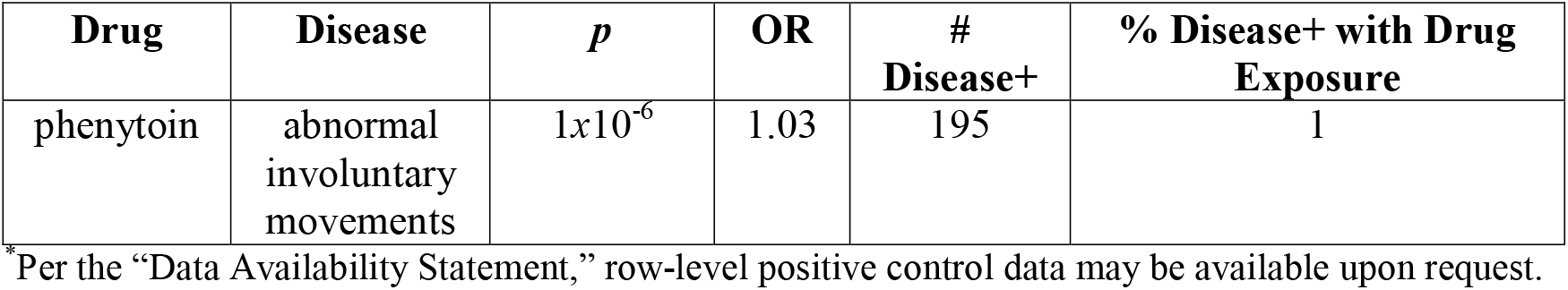
An example MedWAS outcome for a known teratogenic relationship (fetal phenytoin intoxication and chorea) shows statistical significance across several mother-baby pairs, as we expected. A collection of 21 similar results across 10 known teratogens supports our claim of proof-of-concept for our approach.

| **Drug** | **Disease** | ***p*** | **OR** | **#**<br>**Disease+** | **% Disease+ with Drug**<br>**Exposure** |
| --- | --- | --- | --- | --- | --- |
| phenytoin | abnormal<br>involuntary<br>movements | $1 \times 10^{-6}$ | 1.03 | 195 | 1 |

<sup>\*</sup>Per the “Data Availability Statement,” row-level positive control data may be available upon request.

We considered Bonferroni significance a soft constraint, given increasing consensus that purely statistical significance does not directly imply biological significance—especially in the context of holistic approaches like PheWAS^55^. Instead, we maintained signals with significant *p*-values at a baseline of 95% confidence even if they did not demonstrate Bonferroni significance, relying on the other evidentiary filters we describe below to determine their relative importance. This approach to determining signal significance holds in all places in which we discuss “significant” outcomes within this manuscript.

### Top-Ranking Signals

With a list of convincing drug-disease hypotheses, anchored in statistical significance, literature evidence of preclinical and clinical toxicity, the norms of pediatric and obstetric practice, and replicative case series, we identified several classes of drugs with convincing signals of fetal toxicity that we believe warrant further assessment through more structured epidemiological investigations. These demonstration signals demonstrate the power of our MedWAS approach to generate a pliable, hypothesis-generating pipeline for the stimulation of post-market regulatory review programs for drug safety in pregnancy.

The following classes of drugs appeared most significantly linked to clusters of adverse neurological Phecodes diagnosable in the perinatal period, including “spina bifida” (*n* = 219 children), “neural tube defects” (*n* = 242 children), epilepsy and convulsions (*n* = 2,343 children), abnormal (involuntary) movements (*n* = 602 children), (obstructive) sleep apnea (*n* = 1,376 children), and “infantile cerebral palsy” (*n* = 149 children). We present these relationships not solely from statistical results, but from considering the holistic evidence review that we describe above:

- With limiting *p* = 4*×*10^−10^ and OR = 1.03, anti-epileptic drugs (including gabapentin, a drug routinely used off-label^56^, and known toxicants like valproate and topiramate^57^, as described above)
- With limiting *p* = 2*×*10^−7^ and OR = 1.06, psychotropic agents (including alprazolam and other anxiolytic agents, which are often consumed by pregnant patients but have conflicting safety data on their labels^58^)
- With limiting *p* = 1*×*10^−4^ and OR = 1.02, anti-emetic drugs (including ondansetron, which, while numerously studied in relationship to fetal cardiovascular outcomes^59^, is often consumed in the first trimester and features controversial associations to pediatric central nervous system abnormalities^60,61^)
- With limiting *p* = 8*×*10^−8^ and OR = 1.50, narcotic analgesics^62^ (including fentanyl, which featured > 60% co-incidence rate between maternal drug exposure and detrimental neonatal phenotype and occurred with similar disease links and co-incidence rates to the opiate antagonist naloxone)
- With limiting *p* = 4*×*10^−3^ and OR = 1.83, anti-cancer drugs (including tamoxifen, a drug with few uses among pregnant women who choose not to terminate their pregnancies upon a cancer diagnosis—despite its narrow therapeutic index, the drug does not feature a contraindication for pregnancy on its label^63^)

Our teratology quantitative structure-activity relationship model we describe in “Introduction”^23^ concorded with our present analysis of drugs containing fluoroquinolone and azetidinone motifs, providing us with an additional layer of validative evidence review in support of the performance of our process.

### Overall Limitations and Next Steps

Our results demonstrate that systematic assessment of the pharmacological determinants of pregnancy outcomes is possible with simple methods of RWE synthesis that repurpose information routinely collected from primary care and are sufficiently flexible to accommodate direct input from the clinical stakeholders who provide care to pregnant patients and their newborn children. In this regard, our process presents the importance of complementing quantitative methods with qualitative evidence, as much of the contextual knowledge on obstetric prescriptive practice and pediatric disease assessment remains unavailable in structured databases. This combination of ML and consensus prioritization among human users for accurate outcomes analysis is archetypal of PheWAS and GWAS approaches, as many previous publications affirm^21,35,64^.

Our signals present exciting opportunities for confirmation and further interrogation through mechanistic models of human development, as well as for more rigorous evaluation through regulatory-facing program development^65–67^. This expansion is facilitated by the availability of an ontology of medical record numbers for patients with each drug exposure and each outcome that we tested, facilitating review of individual EHRs to confirm true incidence of prescription and disease, as well as to understand confounding variables within the natural history of patients’ care that our quality control system did not consider but may otherwise explain disease signals. These chart reviews are important and must be undertaken rigorously (e.g., through a repeated random sampling approach) for each drug class in which there is interest in deeper study. In this way, continuously integrating knowledge about the clinical implementation of implicated agents and the manifestations of their related diseases will allow for further specification of our hypothesis generation platform in the more probative research that we have planned in the future.

We again affirm that the goal of our research was the development of an enterprise-wide, hypothesis-generating pipeline of drug safety signals using clinical trial emulation. This work does not aim to identify malpractice and does not comprise clinical guidance on prescriptive behavior for pregnant patients.

Despite this orientation and the advantages of our approach, our methods have important limitations that can also spark new research questions. Beyond the randomization we describe above, ontological barriers prevented us from executing PS to explicitly balance our cohorts before attempting MedWAS for the drug-disease inference within closely matched sub-groups. We considered alignment of maternal morbidity to the Charlson comorbidity index^68^ and application of the superficial method of PS developed by Choi *et al*. for PheWAS-empowered drug development studies^69^, to match mothers with similar baseline medical and demographic histories for comparison through MedWAS. While, if successful, this approach could have increased the resiliency of our analyses to confounding from variables extraneous to the prescription of the drug specified for each trial, we realized that the number of maternal-fetal linkages from a single academic medical center like ours is too low to achieve the maximal level of controlling *in situ*. While ∼100K EHRs is a moderately-large data set for implementation of the present research—and represents the data captured from a large, productive medical center— this project demonstrated that execution of our methods with automated controls for confounding by maternal disease history and patient demography requires access to larger databases to prevent attrition of all comparable patient records. Though we could not employ PS *in situ*, as we had originally hoped, we believe that the evidence synthesis workflow we developed—along with the availability of manual patient chart review modules alongside MedWAS—successfully helped us to address the effects of these potentially confounding variables through our signal vetting and prioritization procedure. In future research, we hope to access larger administrative databases of patient records, so we may better integrate PS into our quantitative process. This access could also allow facilitate testing against more discrete representations of neonatal phenotypes than those encoded by Phecodes.

We affirm throughout this manuscript that a central challenge to studying pregnancy and its outcomes with EHRs is defining the period of gestation. Many EHR systems rely on a “pregnancy flag,” encoding, on the backend, a binary representation of pregnancy status^70^. This flag is problematic^71,72^, as we have noticed in our EHR system that it often triggers by elevation in a patient’s body mass index. Therein, reduced precision from the available marker means that inferential approaches to defining the period of pregnancy are necessary to layer other study elements, such as identifying a patient’s medication history during gestation. Arithmetic approaches—such as subtracting 40 weeks from a patient’s delivery date documented on a labor and delivery form to estimate conception date—are possible for first-pass estimation of gestational period, but they rely on low missingness in delivery date information within a candidate EHR data set. Contrastingly, as we describe in “Methods,” we found that an inferential approach to predicting the first date of gestation is an appropriate pathway to addressing pregnancy identification, as data missingness in the extraction of delivery date from the provider-facing EHR to institutionally-maintained databases for secondary use is surprisingly significant. Our approach is also more accurate than the arithmetic approach, as it relies on multiple validated signals of obstetric care. We consider this approach more parsimonious than one of systematic data imputation followed by arithmetic determination, and we affirm that detailed informatics of pregnancy determination in the EHR lie outside the scope of the present study. Similarly, we are unaware of a row-level data source on the natural history of pregnancy that does not present such ontological limitations or that does not require statistical approaches to defining gestational time. Our approach is sufficiently simple to work across other data sets aligned to the Observational Medical Outcomes Partnership common data model (CDM)^73^, while enabling our analysts to readily reproduce our phenotyping for future experiments at our institution, given the approach’s training on our EHR data.

Also, by design, our study evaluates perinatal outcomes, as testing relationships between *in utero* drug exposures and phenotypes at prolonged stages of the pediatric life course remains highly difficult due to the accumulation of potentially confounding etiologies during the natural history of childhood^74^. Therefore, we do not consider the boundaries of our phenotyping capabilities as a limitation of our approach; however, we believe that quantifying the extent to which drug exposures during pregnancy can create lifelong disabilities is a significant question. Addressing this question remains a “grand challenge” in the fields of pharmacoepidemiology and life course research and therefore warrants the creation of data management infrastructure that is more capable of reliably capturing patients’ childhood progressions through a collection of systems more diverse than EHRs^75^. We affirm that our decision to implement MedWAS across all pediatric outcomes, with downstream filtration of results to only perinatal outcomes, allowed us to accomplish our goal of discovering potentially iatrogenic etiologies for birth defects, while also allowing us to prospectively harness our data for future studies of prenatal determinants of adverse health outcomes that manifest later in childhood.

In keeping with the results of most PheWAS studies, we are aware that the signals generated from this platform are potentially controversial^76^ and that despite our attempts to integrate multiple streams of clinical, statistical, biological, and archival evidence with manual EHR review, several of the hypotheses we generated may be explained by non-pharmacological factors. We believe, however, that the strength of our platform is in the identification of priority areas for post-market review of drug use during pregnancy that is anchored in simple methods of RWE, that is sufficiently robust to accommodate the diversity of maternal drugs and perinatal diseases that naturally manifest in a large health system, and that is sufficiently parsimonious to allow for process replication in other health enterprises. We consider that the limited preconditioning necessary for execution of our approach makes it pacakageable, and that the qualitative aspects of our study design allow us to engage necessary clinical stakeholders for drug review more closely than further automated approaches might.

We envision that this work will allow us to partner with regulators of consumer drug products to develop new programs that review post-market drug use among pregnant patients. Presently, we are working with colleagues at FDA to expand our model on larger and more diverse data sets and to integrate new evidence streams into our evaluation process, so we may achieve this goal of regulatory impact, while enhancing the analytical power and extent of trial emulation inherent to our approach. We consider the engagement of regulatory stakeholders as a very positive outcome of this research, which reinforces its implementabilty and provides us with an avenue to attempt the next steps we describe above. Furthermore, as part of a bench-to-bedside initiative to generate more accurate signals of drug safety in the regulatory evaluation of drug products potentially prescriptible to pregnant people, we are currently developing organotypic models of the human placenta^77^ and developing brain^78^ that can allow us to validate our most convincing MedWAS signals on a mechanistic basis.

## Conclusions

The systematic exclusion of pregnant people from RCTs remains common practice in the development and regulation of most consumer drug products; while this approach protects expectant mothers and their unborn children from potentially untoward side effects of experimental drug candidates, it translates to a persistent lack of clinical-grade information on the safety of most drug products employed at the point of care. Therefore, given the bulk of drugs for which it was unethical to include pregnant patients in RCTs but through which this population now receives therapeutic benefit, systematic and robust approaches to post-market surveillance of drug safety in pregnancy are important to ensure equipoise in ethics and health equity for this vulnerable patient population. The use of high-dimensional RWE in this space allows for efficient and representative post-approval analytical processes, as human data routinely generated from primary care may be repurposed in strategic ways to identify areas warranting further assessment for regulatory intervention and quality assurance.

The research that we present in this manuscript proposes and demonstrates the development and implementation of such an approach to safety assessment. Through analysis of EHRs from a high-volume clinical enterprise, we present a workflow of mixed-methods evidence synthesis from clinical trial emulation and the engagement of key stakeholders in the delivery of maternal-fetal care. Our approach allows for efficient, high-level identification of drug classes that warrant further consideration for regulatory program development through more structured, epidemiological methods. We are now constructing these downstream validation programs.

In this manuscript, we also unpack the paradox that while regulatory-facing review of EHR data is one of very few ways to systematically access data on human drug response during pregnancy, the EHR requires important modifications to increase its amenability for such studies. Herein, while our signal identification and validation efforts have important limitations, we believe that our workflow can generate, at scale, a library of interesting and data-driven hypotheses on drug safety risks in pregnancy. We also value this workflow’s emphasis on hypothesis prioritization and the engagement of clinical providers in discussions on the need for enhanced labelling of pharmaceutical products. We believe the simplicity of our process makes it sufficiently pliable to implement at other health systems with the same CDM underlying their EHRs, which we hope will further advance our understanding of the robustness of our approach.

If we can expand our approach and further assess its robustness through the guidance of our regulatory collaborators, we believe that the research we present here could have important applications to enhancing the continued safety assessment of many drugs commonly consumed by the pregnant public. This goal holds immense translational potential.

## Abbreviations

ATC: Anatomical Therapeutic Chemical
CDM: common data model
CDS: clinical decision support
EHR: electronic health record
FDA: United States Food and Drug Administration
ICD: International Classification of Disease
IRB: institutional review board
KMCI: KnowledgeMap concept identifier
MedWAS: medication history-wide association study
ML: machine-learning
NCATS: National Center for Advancing Translational Sciences
NICHD: National Institute of Child Health and Development
NIH: United States National Institutes of Health
NLP: natural language processing
OCM: Office of Contracts Management
OR: odds ratio
OTC: over-the-counter
PheWAS: phenome-wide association study
PS: propensity scoring
RCT: randomized, controlled trial
RD: Research Derivative
RWE: real-world evidence
VUMC: Vanderbilt University Medical Center

## Author Contributions

APC led the study team. APC, XN, LMB, ESL, RRL, and JAG designed the platform described in this manuscript. XN and LMB, as the data analysts for this project, determined gestational ages, executed MedWAS and its associated sensitivity analyses, and evaluated the compatibility of PS matching with our data source. APC, EAG, ESL, RRL, and JAG developed the evidence synthesis procedure for MedWAS validation and signal prioritization. EAG, SLV, and DMA provided clinical consultations on MedWAS outcomes. ESL provided neuroscientific consultation on MedWAS outcomes, and RRL provided pharmacological consultation. APC drafted this manuscript for the study team’s review, and XN drafted the expanded methods section included in the supplementary material that accompanies the manuscript. APC created the figures in this manuscript. All co-authors were consulted to review this manuscript.

## Acknowledgements

APC, DMA, and EAG acknowledge support from award R21HD105304 from National Institute of Child Health and Development (NICHD) of NIH. APC, DMA, and ESL also acknowledge relevant funding from award U54TR02243-02 from National Center for Advancing Translational Sciences (NCATS) of NIH. APC and DMA are additionally funded for a study of therapeutic index in pregnancy by award N009367701 from Rainwater Charitable Foundation. SLV was supported by IRSA #1015006 from the Burroughs Wellcome Fund.

We thank Noel Southall, PhD, Team Leader in NCATS’s Division of Preclinical Innovation and the NCATS-FDA interagency initiative, for his advice on the framing and applications of the work we describe in this manuscript. We also thank Andrew Beam, PhD at Harvard T.H. Chan School of Public Health for his feedback on study design and results interpretation and Meghan Vance at VUMC for her help with developing Figure 3.

We acknowledge that the perspectives we present in this manuscript are solely the authors’; they do not necessarily represent the official views of NCATS, NICHD, or any other organization.

