## Supplementary Information for "Clinical trial emulation can identify new opportunities to enhance the regulation of drug safety in pregnancy"

| David M. Aronoff, MD, FIDSA, FAAM  Indiana University School of Medicine,  Emerson Hall 305,  545 Barnhill Drive,  Indianapolis, Indiana 46202,  United States of America   Tel. (desk): +1-317-274-8438  Tel. (fax): +1-317-274-1437 | Anup P. Challa, BEng, MS, EIT  Vanderbilt University and Medical Center,  PMB 350969,  2301 Vanderbilt Place,  Nashville, Tennessee 37203,  United States of America   Tel. (desk): +1-615-875-0085  Tel. (fax): +1-615-936-8545 |
| --- | --- |

**Table of Contents**

*Item*	*Page*

Introduction………………………………………………………………………………………..3

Expanded Methods Cookbook………………………………………………………….………....4

I. Defining Period of Gestation……………………………………………………………...4
II. Collecting Mothers’ Medication Data during Gestation…………………………..……...7

References………………………………………………………………………………………..21

| **Introduction** |
| --- |

We provide the following expanded methods and characterization of our mother-baby cohorts with hope that these data enhance the reproducibility of the results that we detail in our manuscript. Though, as we describe in our manuscript, regulations that protect the security of the patient data that underlie our MedWAS prevent disclosure of the associated outcomes, we hope that this supplement conveys the pliability of our approach. For extraneous data requests, please contact the Corresponding Authors at the email addresses provided above.

**Expanded Methods**

*I. Defining Period of Gestation*

Consider a data set in the form of a list of mother and baby link/pair with accompanying demographic data for each pair.

Use ICD-10 encoding of weeks of gestation Z3A* to determine mothers’ day 1 gestation and period of gestation (the asterisk symbol after the primary code, Z3A, means to include all of secondary codes after Z3A. Detailed information for Z3A and its secondary codes are accessible here: <https://www.icd10data.com/ICD10CM/Codes/Z00-Z99/Z30-Z39/Z3A->.

1. For mothers who got ICD-10 codes for gestation weeks large than 8 weeks and less than 42 weeks (From Z3A.08 to Z3A.42):
   a. Search against the list of mother-baby link to identify mothers who got ICD-10 codes from Z3A.08 to Z3A.42.
   b. Get mothers’ earliest and latest coding dates for those codes for each pair of mother and baby pair.
   c. Calculate the days between the latest and earliest ICD-10 code dates and if the calculated days is less than 250 days, derive the 1^st^ day of mothers’ gestation by subtracting the latest coding date with the days calculated from the weeks of gestation multiplied by 7 days.
   d. Calculate the period of mothers’ gestation by using the first day of mothers’ gestation derived from the latest ICD code date and babies’ birth date.
   e. For the calculated days from step c > 250 days, derive the first day of mothers’ gestation by subtracting the earliest coding date with days calculated from the weeks of gestation multiplied by 7 days.
   f. Calculate the period of mothers’ gestation by using the first day of mothers’ gestation derived from the earliest ICD code date and babies’ birth dates.
2. For mothers who got ICD-10, Z3A.01, less than 8 weeks:
a. Search against the list of mother-baby pairs to identify mothers who got Z3A.01 less than 8 weeks of gestation.
b. Get mothers’ earliest and latest coding dates for Z3A.01.
c. Calculate the days between earliest and latest coding dates for Z3A.01.
d. Keep mothers with 0 < the days between earliest and latest coding dates for Z3A.01 < 56 days.
e. Determine or keep mothers’ day 1 gestation by subtracting the latest coding date with 28 days or 4 weeks if the days (period of gestation) calculated from babies’ birth dates to mothers’ day 1 gestation is between 150 and 295 days.
3. For mothers who got ICD-10 larger than, Z3A.49, 42 weeks:
a. Search against the list of mother-baby pairs left over from Step 2 to identify mothers who got Z3A.49, large than 42 weeks gestation of pregnancy.
b. Get mothers’ latest coding dates for Z3A.49.
c. Estimate the first day of gestation by subtracting the latest coding date with 42 weeks multiplied by 7 plus an extra 4 days.
d. Calculate mothers’ period of gestation by using the first day of gestation and babies’ date of birth and only keep the calculated period of gestations larger than 295 and less than 320 days.

Combine the data sets from steps 1, 2, and 3, and denote the resulting data as “mother-baby_icd10.”

Now, use the Systemized Nomenclature of Medicine (SNOMED)^1^ to determine mothers’ day 1 gestation and period of gestation. To implement this approach, EHR data should be stored under the Observational Medical Outcomes Partnership (OMOP) common data model^2^.

1. Prepare the list of mother-baby pairs whose day 1 and period of gestation were not determined using above ICD-10 code approach.
2. Collect a list of concept_id from table, CONCEPT, by specifying the field, concept_name having the keywords, ’gestation period’ and the key word, ‘weeks’ as the last word in the value of each concept_name. Also, the field vocabulary_id needs to be specified with “SNOMED” to define the key words search is limited to be from SNOMED.
3. Collect mothers’ IDs, SNOMED codes, condition_source_value, condition_start_date in table CONDITION_OCCURRENCE and then merge them with concept IDs from step 2.
4. Group the records based on mothers’ ID and derive the earliest and latest dates for a mother who got those SNOMED codes.
5. Keep those mothers whose days between earliest and latest date of SNOMED is less than 280 days but larger than 0 days.
6. Deduce mothers’ day 1 of gestation by using the latest date of getting SNOMED to subtract the days calculated from the weeks of gestation multiplied by 7. Then, calculate the period of mothers’ gestation by using babies’ birth dates and the derived mothers’ day 1 of gestation.
7. Also, keep those entries with mothers whose days between earliest and latest date of getting mapped SNOMED is large than 280 days.
8. For mothers coming from step 7, deduce mothers’ day 1 gestation by using the date of earliest getting SNOMED to subtract the days calculated from the weeks of gestation multiplied by 7, and then calculate the period of mothers’ gestation by using babies’ birth date and the derived mothers’ day 1 gestation.
9. Pool datasets from steps 6 and 8 and denote it as “mother-babysnomed.”

Now, use ICD-9 and Current Procedural Terminology (CPT) billing codes to determine mothers’ day 1 gestation and period of gestation.

1. Prepare mother-baby dataset which wasn’t determined by the strategies of using ICD-10 and SNOMED.
2. Search above dataset with ICD-9 codes, v72.42, v22.0, and v22.1 to create a cohort, from which mothers are currently getting positive test results (v72.42) for normal pregnancy (v22.0 and v22.1).
3. Collect data from the cohort created in step 2 by keeping only these mothers who also got CPT codes, '76801' and '76802'.
4. Derive mothers’ day 1 gestation by using the date of CPT codes minus 98 days.
5. Calculate the period of mothers’ gestation using mothers’ day1 gestation and babies’ birth date and then only keep the period of mothers’ gestation is between 260 and 310 days.
6. Name the dataset as “mother-baby_ICD9_CPT”.

Finally, pool the 3 datasets, mother-baby_icd10, mother-baby_snomed, and mother-baby_ICD9_CPT to be the final dataset, which is used for rest of MedWAS analysis.

*II. Collecting Mothers’ Medication Data during Gestation*

Consider the defined mother-baby dataset from section I, which is a list of mother and baby pairs with their demographic information, day 1 gestation, and the estimated period of gestation in days. Here, we need to prepare medication data during the estimated period of mothers’ gestation and assume that all EHR data for replicative analyses are implemented using OMOP schema.

1. Specify medication data collection is based on the drug ingredient by limiting CONCEPT_CLASS_ID=’ Ingredient’.
2. Collect concept_id and concept_name (drug_ingred) using SQL code like that below.

SELECT DESCENDANT_CONCEPT_ID AS CONCEPT_ID,CONCEPT_NAME AS DRUG_INGRED FROM

(select * from (

SELECT DISTINCT CONCEPT_ID,CONCEPT_NAME,CONCEPT_ID AS ANCESTOR_CONCEPT_ID,CONCEPT_CODE FROM (

select * from concept where

upper(domain_id)like upper('drug%')and

UPPER(CONCEPT_CLASS_id)LIKE UPPER('INGREDIENT%'))D_INGREDIENT)P

JOIN

CONCEPT_ANCESTOR B

using(ancestor_concept_id))PC_J

3. Join or merge patients’ EHR medication data in table, DRUG_EXPOSURE, with drug_ingred (concept_name) using concept_id from step 2.
4. First, collect mothers’ medication data during the period of gestation by merging or joining data from step 3 using mothers’ IDs from the defined mother-baby dataset developed in section I.
5. Second, define the window for collecting mothers’ medication exposure during pregnancy as 60 days before the first day of mothers’ gestation until birth dates.
6. Option #1: to collect mothers’ medication data during the period of gestation as binary data, just assign “1” as drug exposure. Or, option #2: to collect mothers’ gestational medication data as abundance data, cumulatively count medication exposure times.

Prepare a medication exposure data file in the format of binary or abundance, as in the screenshots below (Figures S.1 and S.2).

Note: When dealing with drug’s name, eliminate any none (Aa-Zz and 0-9) characters and replace ‘white space’ with ‘_’ to get final drug names. For example, the drug name, “INFLUENZA VIRUS VACCINE, INACTIVATED A-CALIFORNIA-07-2009 X-179A (H1N1) STRAIN“, was changed to “INFLUENZA_VIRUS_VACCINE_INACTIVATED_A_CALIFORNIA_07_2009_X_179A_H1N1_STRAIN.”

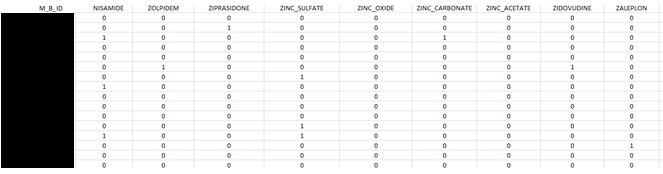
**Figure S.1**: Medication data in binary format (to protect patient privacy, mother-baby linkage identifier is redacted in this display)

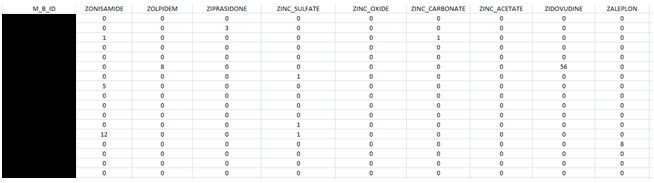
**Figure S.2**: Medication data in abundance format (to protect the security of patients’ data, mother-baby linkage identifier is redacted in this display)

*III. Collecting Children’s Phenotype Data*

Consider the defined mother-baby dataset from section I, which is a list of mother and baby pairs with their demographic information, day 1 gestation, and period of gestation in days. Here, we identify phenotypes for all candidate children and align the phenotypes to Phecodes.

1. The time window to collect babies’ phenotypes is defined as from the date of birth specified to when the babies turn 18 years old.
2. Based on the times defined from step 1, babies’ ICD-9 and ICD-10 codes are collected and stored in a CSV file (per the screenshot Figure S.3, below) using babies’ ID from the defined mother-baby dataset.

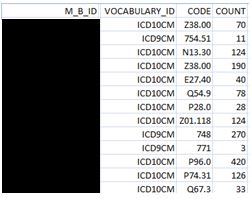

**Figure S.3**: Phenotype data (as ICD-9 or ICD-10 codes) for candidate children—“COUNT” specifies the number of instances of the code in the child’s EHR during the period of observation. To protect the security of patients’ data, the mother-baby linkage identifier is redacted in this display.

3. The collected ICD-9 and ICD-10 codes above are converted to Phecodes (per the screenshot Figure S.4, below) using the PheWAS package download from the following website: <https://www.vumc.org/cpm/center-precision-medicine-blog/phewas-r-package>. The details of converting ICD-9 and ICD-10 codes to Phecode are accessible through the PheWAS package.

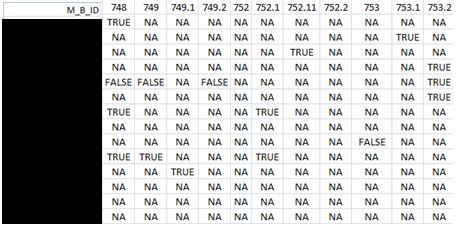

**Figure S.4**: Phenotype data aligned as Phecodes following execution of the PheWAS package—Phecodes converted from the ICD ontology are displayed from the second column onwards. “TRUE” means the baby was phenotyped as case for a MedWAS test, “FALSE” stands for the baby got phenotyped as control for that test, and “NA” denotes that the baby was phenotyped as neither case nor control. To protect the security of patients’ data, the mother-baby linkage identifier is redacted in this display.

*IV. Running MedWAS with the PheWAS Package*

Consider the medication dataset created from section II, which stored in the format of CSV flat file. The phenotype dataset created from section III is stored as an R object following import and execution of the PheWAS R package. Here, we demonstrate an example of running association analysis between the children’s phenotypes and mothers’ medication data in binary format during the period of their gestation.

1. Create a mothers’ medication R object by loading the medication dataset file to R using the command below.

medx_data_bin <- read.csv("final_medx_flat_data_bin.csv",head=T,sep=",")

2. Run MedWAS by looping each drug (from drug 1 to n, where n denotes the number of the drugs available for MedWAS) in medx_data_bin using the command below.

drug_pheno_as1-n <- phewas(phenotype_data, drug1-n_data)

3. Assign phenotype information for each drug-phenotype association output from step 2 by using the command below.

drug_pheno_as1-n <- addPhecodeInfo(drug_pheno_as1-n)

4. Output each drug-phenotype association output as CSV files using command below.

write.csv (drug_pheno_as1-n, "./output_directory/drug_pheno_as1-n,".csv",sep='')

5. Loop each outputted drug_pheno_as1-n.csv to remove any row with the eighth field having the value “A,” and only keep the fields {2,3,5,6,7,8,9,10,11,12,16}. Finally, merge all fields one file, which is the final MedWAS outcome/data file (see screenshot Figure S.5, below)

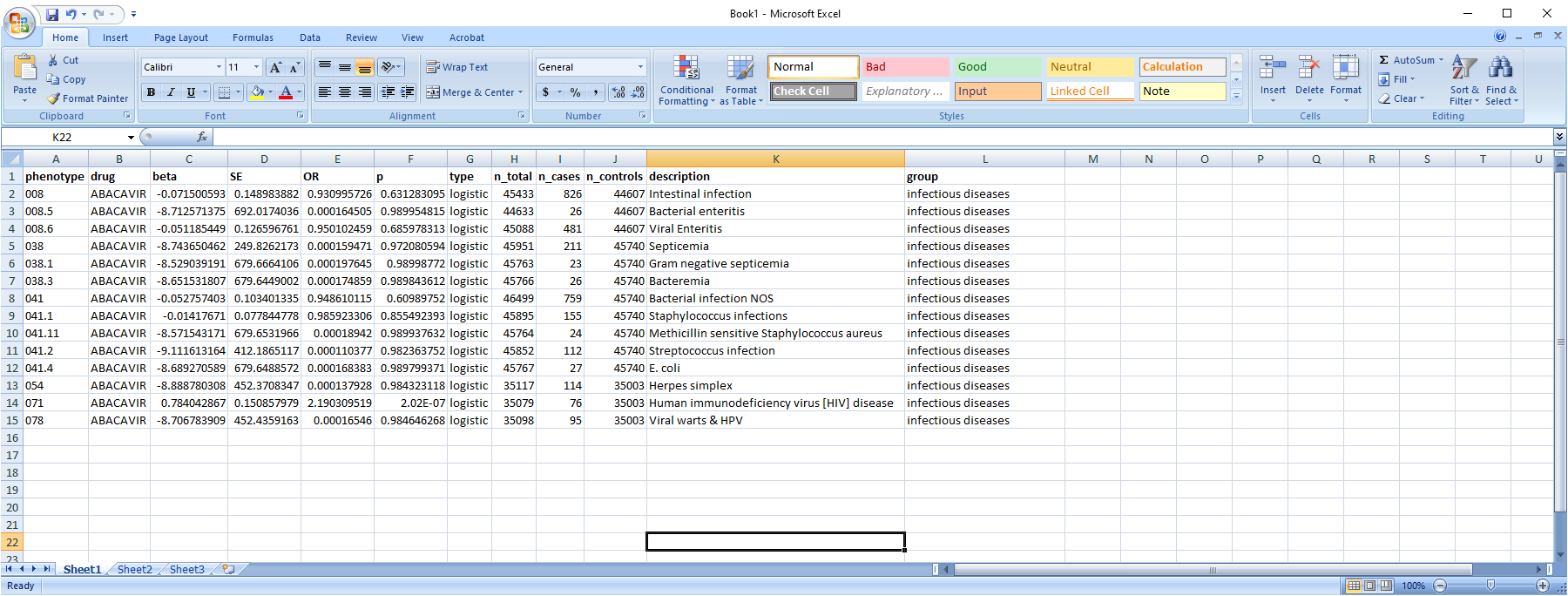

**Figure S.5**: Example instances in an outputted MedWAS data set

6. To count the number of drug exposure and no drug exposure from phenotype and control, write script in your preferred programming language to scan the phenotype data and medication data files for each participating mother-baby pair and then merge medication counts underlying each associated drug with above MedWAS outcome to obtain a harmonized data set.

**Dictionary of Maternal Medication Data**

The following list gives all pharmaceutical and nutraceutical agents documented in pregnant patients’ EHRs, per the medication identification approaches we describe in our manuscript. We filtered drug signals from this raw, holistic list and removed any strings that are components of larger drug names, which we considered artifact from the performance of our NLP tools.

| Probenecid | Neomycin | Granisetron | Titanium |
| --- | --- | --- | --- |
| Atracurium | Enalaprilat | Colistimethate | Ibuprofen |
| Indole | Cycl | Procainamide | Pantoprazole |
| Val | Dexlansoprazole | Regadenoson | SMX |
| Sotalol | Valsartan | Epinephrine | Turmeric |
| Cetylpyridinium | Carbamazepine | Vinorelbine | Prilocaine |
| Ranibizumab | Potassium Iodide | Amprenavir | Platinum |
| Ceftizoxime | Atovaquone | Metyrapone | Dexmedetomidine |
| Levofloxacin | Cevimeline | Sesame oil | Ephedrine |
| Triamterene | Clorazepate | Candida albicans | Selenium Sulfide |
| Hydroxyprogesterone | Ledipasvir | Calcium Carbonate | Folate |
| BCG vaccine | Nizatidine | Oxybutynin | Chlorzoxazone |
| Ceftaroline fosamil | Chloride | Cisplatin | Sulfadiazine |
| Rituximab | Chlorophyll | Zinc oxide | Famciclovir |
| Misoprostol | Benzonatate | AS | Iodide |
| Calcium acetate | Carbon monoxide | Mercaptopurine | Bisacodyl |
| Fluvoxamine | Telmisartan | Black cohosh | Pyrazinamide |
| Lisdexamfetamine | Tenofovir alafenamide | Estradiol valerate | Duloxetine |
| Fosfomycin | Lactobacillus reuteri | Methyl salicylate | Dextromethorphan |
| Phenylephrine | Adalimumab | Naltrexone | Grape |
| Sucralfate | Uric Acid | Omeprazole | Potassium Citrate |
| Amino acids | Enalapril | Pseudoephedrine | Promazine |
| Dutasteride | Linaclotide | Vancomycin | Lemon |
| Memantine | Papaverine | Baclofen | Dantrolene |
| Diphenoxylate | Choline | Perphenazine | Ustekinumab |
| Methylcobalamin | MoCo | Pioglitazone | Opium |
| Pancuronium | Calcio | Arginine | Sucrose |
| Praziquantel | Flur | Dextrose | Carbidopa |
| Gentamicin | Milrinone | Kale | Butalbital |
| Mepivacaine | omycin | Sesame seed | Glucagon |
| Diflucan | Prazosin | Amoxicillin | Triamcinolone |
| Mexiletine | Salmeterol | Flax seed | Cyclosporine |
| Chloral hydrate | Permethrin | Heparin | Sulindac |
| Ferrous asparto glycinate | Mesalamine | Methadone | Desmopressin |
| Aztreonam | Esomeprazole | Nebivolol | Telbivudine |
| Atenolol | Bethanechol | Ketorolac | Asp |
| Ketoconazole | Honey | Danazol | Prednisolone |
| rina | Primrose oil | Silver sulfadiazine | Bimatoprost |
| Iodine | Vitamin B2 | Black walnut | dDAVP |
| Ecgonine | Vitamin B12 | Mafenide | Hydrocortisone valerate |
| Tea tree oil | Irbesartan | Insulin Aspart | Itraconazole |
| Cocoa | Magnesium chloride | Von willebr | Cephradine |
| Imiquimod | Poractant alfa | Chlorthalidone | Bisoprolol |
| Ketamine | DCI | Calciferol | Melatonin |
| Calcium gluconate | Zn | Nitroglycerin | Cod |
| Montelukast | Ferrous fumarate | Cefpodoxime | Magnesium citrate |
| Castor oil | Valproic Acid | UDCA | Cocaine |
| Rizatriptan | Indinavir | Synephrine | Borage oil |
| Omega-3 | Vardenafil | Azathioprine | Cefprozil |
| Medroxyprogesterone acetate | Meclizine | Succinic acid | Pilocarpine |
| Aspart | Phenobarbital | Thiopental | Homatropine |
| Cyclophosphamide | Perazine | Cyanocobalamin | Bupivacaine |
| Carbonyl iron | Beclomethasone dipropionate | Dydrogesterone | Leflunomide |
| Darbepoetin | CD | Escitalopram | Urea |
| Nelfinavir | Niacin | Fentanyl | Hydroxocobalamin |
| Octreotide | Tranylcypromine | BNP | Mefenamic acid |
| Ozone | d-amphetamine | Modafinil | Vanilla |
| Desonide | Adapalene | Air | Hydrocodone |
| Midazolam | Vilanterol | Tetracycline | Infliximab |
| Fluocinolone | Simethicone | Maltose | Omalizumab |
| Fludrocortisone | Cellulose | Paprika | Papaya |
| Corn | Clarithromycin | Chlorpromazine | Magnesium oxide |
| Non | AMP | Ac | Ezetimibe |
| Phentermine | Sevelamer | Ergocalciferol | Enoxaparin |
| Dobutamine | Leucovorin | Barium | Spironolactone |
| Pirbuterol | Fexofenadine | Colace | Donepezil |
| Latanoprost | Rosin | Norgestimate | 5 |
| Methylphenidate | Theophylline | Raltegravir | Lamotrigine |
| Bepotastine | Carbonate | Cinnamon | Timolol |
| Magnesium sulfate | Tramadol | Vitamin E | Dipyridamole |
| Ginseng | Cobalamin | Betaxolol | Corticotropin |
| Zinc | Etanercept | Chlorhexidine | Sulbactam |
| Cloxacillin | Oregano | Succinate | Pea |
| Selegiline | Idarubicin | Lithium citrate | Neostigmine |
| Clonazepam | APAP | Deferasirox | Lurasidone |
| Meloxicam | Darunavir | Eplerenone | Glycerin |
| Citric Acid | Evening primrose oil | Chloroquine | Nitric Oxide |
| Hydroxychloroquine | Rotavirus Vaccine | Ranitidine | ASC |
| Lutein | AVAC | Fenofibrate | Water |
| Naproxen | Famotidine | Desipramine | Aripiprazole |
| Nortriptyline | Nitrogen | Metronidazole | Metolazone |
| Fish oil | Formoterol | Astaxanthin | Ethynodiol diacetate |
| Milk of magnesia | Cefotaxime | Iohexol | Acetate |
| Canagliflozin | Vinegar | Pitavastatin | Hypromellose |
| Daunorubicin | Metoclopramide | pC | Calcitriol |
| Ginkgo biloba | Risperidone | Vitamin A | Felodipine |
| Aloe | Gemfibrozil | Antipyrine | Erythromycin |
| Tolnaftate | Vanadium | Loratadine | DM |
| Flecainide | Sumatriptan | Fosamprenavir | Hydrocortisone acetate |
| Voriconazole | Salsalate | Fluorouracil | osterone cypionate |
| Anastrozole | Aminocaproic Acid | Fluticasone | Petrolatum |
| Budesonide | Haloperidol | Hydroxyzine | Losartan |
| Mometasone | Vitamin B1 | Levonorgestrel | Diatrizoate |
| Chromium picolinate | Clozapine | Butamben | Ursodiol |
| Vincristine | Butoconazole | Camphor | Zonisamide |
| Ethyl chloride | Leuprolide | Drospirenone | Atropine |
| Cherry | Cauliflower | Cortisone acetate | Clopidogrel |
| Mineral oil | Cadmium | Riboflavin | age |
| Desvenlafaxine | Clobetasol | Lithium carbonate | Clomipramine |
| Orange | Methotrexate | Doxepin | Econazole |
| Alprostadil | Pramipexole | Codeine | Capsaicin |
| Micafungin | Primidone | Ciprofloxacin | Cranberry |
| Orphenadrine | Indapamide | Tioconazole | Mebendazole |
| Resveratrol | Zinc sulfate | Mometasone furoate | Oxymetazoline |
| Colestipol | Lacosamide | Dexamethasone | Diazepam |
| GnRH | Potassium bicarbonate | Lanolin | Bacitracin |
| BIA | Natalizumab | Tinidazole | Carboplatin |
| Isometheptene | Cyclobenzaprine | Prednisone acetate | Amber |
| Progesterone | Cefoxitin | Biotin | Amlodipine |
| NAC | Sulfasalazine | Phenytoin | Psyllium |
| Clove | Bromocriptine | Tetanus toxoid | Nicardipine |
| Ethosuximide | Everolimus | KCl | Tamoxifen |
| Povidone | Pomegranate | Phosphate | Conjugated estrogens |
| Glu | Sofosbuvir | Asenapine | Etonogestrel |
| Pentoxifylline | Omega-3 fatty acids | Hydrocortisone butyrate | Rabeprazole |
| Trandolapril | Ketoprofen | Nitrate | osterone |
| Sodium fluoride | Pindolol | Temazepam | Ascorbic acid |
| Calcium | Ziprasidone | Eszopiclone | Fluconazole |
| Trimethobenzamide | elate | Pramoxine | Lactose |
| Cilostazol | Garlic | Pyridoxine | Terbinafine |
| Pyrethrins | FTP | Butorphanol | Amitriptyline |
| Inositol | Beta carotene | Silver nitrate | Alprazolam |
| Methylprednisolone | Royal jelly | Apple | Quinine |
| Clobazam | Cocoa butter | Norepinephrine | Minocycline |
| Dornase alfa | Hydrochlorothiazide | iol | Bumetanide |
| Phloroglucin | Sertraline | Etodolac | Abacavir |
| Mefloquine | Cortisone | Orlistat | Droperidol |
| Acetylcysteine | Naloxone | Ondansetron | Argatroban |
| Coconut | Carrot | Atazanavir | Pepsin |
| Erythropoietin | Hydrogen peroxide | Isosorbide | DHA |
| Albuterol | Rice | Carbamide | Pentazocine |
| Venlafaxine | Bromfenac | Estradiol | Valerian |
| Squash | Dopamine | Copper | Vitamin C |
| Rilpivirine | Isotretinoin | Thyroglobulin | DCA |
| Pearl | Sodium lauryl sulfate | Etravirine | Carboprost tromethamine |
| Bupropion | Tropicamide | Propylene glycol | Zidovudine |
| Fluticasone propionate | Kidney bean | Bivalirudin | Ceftriaxone |
| Glucose | Naratriptan | Benzodiazepine | Fluorometholone |
| Hetastarch | Ammonia | Felbamate | Levocetirizine |
| Cetirizine | Metaxalone | Nor | Talopram |
| Trihexyphenidyl | Bean | Mycophenolic acid | Aldosterone |
| Insulin Lispro | Tretinoin | Benzatropine | Caffeine |
| Rocuronium | T3 | Thrombin | Fosinopril |
| Dolutegravir | Celecoxib | Digoxin | Lovastatin |
| Collagenase clostridium histolyticum | PAS | Gelatin | COP |
| Cefaclor | Docusate | Meperidine | Clobetasol propionate |
| Carvedilol | Lansoprazole | Dimenhydrinate | Silver |
| Flavoxate | Pineapple | Fluticasone furoate | Tazobactam |
| Olive oil | Flunisolide | Amiodarone | MPA |
| Ritonavir | Oss | Trospium | Tobramycin |
| Alteplase | Theanine | Alfuzosin | Grapefruit |
| Topiramate | Neon | Norelgestromin | Abatacept |
| Ib | Botulinum Toxin Type B | Finasteride | Acetamide |
| Carbamide peroxide | Nifedipine | Fluorescein | Doxorubicin |
| Ursodeoxycholic acid | Chlorothiazide | Retinol | Nitazoxanide |
| Sodium oxybate | Witch hazel | Imipramine | Wheat bran |
| Acitretin | Pentamidine | Human insulin | Methimazole |
| Ferrous bisglycinate | Buprenorphine | Date | Cholecalciferol |
| Ampicillin | Sulfamethoxazole | Maprotiline | Amantadine |
| Chloroprocaine | Olopatadine | Oyster | Betamethasone |
| Etomidate | Epinastine | Glycine | Ofloxacin |
| Levobunolol | Lactulose | Cimetidine | Sodium Citrate |
| Ciclopirox | Doripenem | Terbutaline | Paroxetine |
| Certolizumab pegol | Rocephin | Curcumin | Tranexamic acid |
| Cisatracurium | Tetrofosmin | Isosorbide dinitrate | Polycarbophil |
| Glutathione | Tapentadol | Terconazole | Calcium Citrate |
| Calcium Chloride | Chlorphenoxamine | Gabapentin | Cod liver oil |
| Inosine | Vitamin D2 | Sage | Flumazenil |
| Cicl | Metformin | Azithromycin | Methyldopa |
| Darbepoetin alfa | Hyaluronidase | Boric acid | Diltiazem |
| Chondroitin sulfate | Lopinavir | Zinc acetate | Lisinopril |
| Lipoic Acid | Exemestane | Ginger | Isoniazid |
| Cholic Acid | Rifapentine | Hyaluronate | Ferric subsulfate |
| Nefazodone | Tacrolimus | Moxifloxacin | Serine |
| Peppermint oil | Sterile water | Folic Acid | Mango |
| Chloroxylenol | Clonidine | Alendronate | Mupirocin |
| Triazolam | Loperamide | Coconut oil | Dextroamphetamine |
| Okra | Tromethamine | Cysteine | Sodium bicarbonate |
| Clavulanate | tPA | Amphetamine | Labetalol |
| Fructose | Methocarbamol | Colchicine | Carbon dioxide |
| Halothane | Almond | Brimonidine | Polyvinyl alcohol |
| Senna | Bevacizumab | axin | Thiamine |
| Levodopa | Desogestrel | Mannitol | Magnesium hydroxide |
| Sitagliptin | Etiracetam | Salicylic acid | Magnesia |
| Valganciclovir | Selenium | Colistin | Aluminum hydroxide |
| Benzocaine | Simvastatin | Galantamine | Minoxidil |
| Dolasetron | Ferumoxytol | Vitamin B6 | Protein C |
| Fosaprepitant | Belladonna | Racepinephrine | Ramipril |
| Talc | Aprepitant | Proparacaine | Rutin |
| Trypsin | Fondaparinux | Nitrofurantoin | Levetiracetam |
| Tolterodine | Insulin Detemir | Ceftazidime | Oxygen |
| Estriol | Nafcillin | Doxycycline | Aluminum chloride |
| Trazodone | Hydromorphone | Remifentanil | Deferoxamine |
| Acyclovir | Histamine | Podofilox | Polymyxin B Sulfate |
| Dextran | Propranolol | Propofol | Vedolizumab |
| Hydroxyurea | Dicyclomine | Vitamin D | Uracil |
| Urethane | Oxazepam | Vitamin D3 | Fluoxetine |
| Glimepiride | Nystatin | IVIg | Tamsulosin |
| Dapsone | Nicotine | Nabumetone | IMD |
| Lactic Acid | Lorazepam | Miconazole | Protamine |
| Propafenone | Rosuvastatin | Potassium gluconate | Promethazine |
| Sirolimus | Tenofovir | Alcohol | Tizanidine |
| Oseltamivir | Adenosine | Scopolamine | Tenofovir disoproxil |
| Meropenem | Terazosin | Proguanil | Procaine |
| Chamomile | Acarbose | Vecuronium | Pyrimethamine |
| Zopiclone | Rivastigmine | Cholestyramine | Benzoyl peroxide |
| Pyrantel | Fluocinonide | Norgestrel | Dimethicone |
| 6 MP | Lamivudine | Azelaic acid | Dorzolamide |
| Milk thistle | Methylcellulose | Zolpidem | Pyridoxal |
| Hydralazine | Perflutren | Factor VIII | Calcium polycarbophil |
| Propylthiouracil | Hyoscyamine | Tetracaine | Morphine |
| Raspberry | Deoxycholic Acid | Sulfacetamide | Cefepime |
| OMO | Bexarotene | Verapamil | Piperacillin |
| Cyproheptadine | Ethanol | Tyramine | Saquinavir |
| Fluocinolone acetonide | Clotrimazole | Methylnaltrexone | Ferric Carboxymaltose |
| Pregabalin | Oxycodone | Letrozole | Atomoxetine |
| Buspirone | Lindane | Methyclothiazide | Cucumber |
| Oxcarbazepine | Ertapenem | Chicken | Pantothenic acid |
| Metaraminol | Ethambutol | Cefdinir | Tiotropium |
| Piperazine | osterone enanthate | Dicloxacillin | Trimethoprim |
| Cefuroxime | Nalbuphine | Albendazole | Doxylamine |
| Benazepril | Detemir | Pravastatin | Iron Dextran |
| Ferrous gluconate | Meprobamate | Glucosamine | Ammonium lactate |
| Linezolid | PAT | Fosphenytoin | Cytarabine |
| Al | Zeaxanthin | Clidinium | Efavirenz |
| Dofetilide | Liothyronine | Gadobutrol | Belimumab |
| Fluvastatin | Captopril | Gold | Epoprostenol |
| Manganese | Peanut | Amiloride | Dronabinol |
| ATT | Brompheniramine | Entacapone | PCA |
| Doxazosin | Ropinirole | Methylosterone | Acebutolol |
| Ivermectin | Glipizide | Nitrous oxide | Chlordiazepoxide |
| Sufentanil | Isosorbide Mononitrate | Prednisone | Alclometasone |
| Diflorasone | Phenazopyridine | Pertuzumab | MCC |
| Pyridostigmine | Acetaminophen | Sodium Chloride | Influenza A virus |
| Sulfate | Hydrocortisone | Sildenafil | Hydroquinone |
| PV | Insulin Glargine | Rabies immune globulin | Malic Acid |
| Phenol | Polyethylene glycol | Diphenhydramine | Carisoprodol |
| Molybdenum | Oxymorphone | Sugammadex | Gadoxetate |
| Peanut oil | Levothyroxine | Aminophylline | Levocarnitine |
| Barbital | Mirtazapine | Lubiprostone | Hepatitis B Vaccine (Recombinant) |
| Benzoylecgonine | Gadolinium | Ganciclovir | Ipratropium |
| Potassium Chloride | Clindamycin | Magnesium gluconate | Glyburide |
| Barium sulfate | Allopurinol | Paclitaxel | Guaifenesin |
| Daptomycin | Magnesium | TMP | Desoximetasone |
| Sennosides | Salbutamol | PPD | Cefazolin |
| Furosemide | Metoprolol | Acetic acid | Esmolol |
| Echinacea | Mycophenolate | Ubiquinol | Inulin |
| Vasopressin | quinol | Cephalexin | Pancrelipase |
| Cefixime | Sodium acetate | Cyclopentolate | Pork |
| Pheniramine | Atorvastatin | Quinapril | Salmon |
| Midodrine | Amphotericin B | Hepatitis A Vaccine | INH |
| Zaleplon | Mifepristone | Hydroxide | Olanzapine |
| Mycophenolate mofetil | Nadolol | ione | Oxytocin |
| Domperidone | Glycolic acid | Ropivacaine | Lidocaine |
| Hydroxyprogesterone caproate | Carbazochrome | Guanfacine | Docetaxel |
| Oxacillin | Quetiapine | Prothrombin | Elm |
| Balsalazide | Acetazolamide | Azelastine | Isoxsuprine |
| Citalopram | Cabergoline | Methenamine | Fluphenazine |
| Phencyclidine | Polystyrene sulfonate | Rifabutin | Botulinum Toxin Type A |
| Iodoform | Potassium acetate | Prochlorperazine | Spiramycin |
| Ribavirin | Iron | 5- | Phloroglucinol |
| Diclofenac | Succinylcholine | Insulin Human | Bismuth Subsalicylate |
| Nevirapine | Potassium | Warfarin | Phosphatidyl serine |
| Aspirin | Granisetron | Emtricitabine |  |

**Cohort Demographics**

Below (Table S.1), we provide summary demographic data about our cohort of mothers and infants. The format of Table S.1 aligns with that mandated by the United States Public Health Service (PHS) for reporting biological sex, race, and ethnicity^3^.

**Table S.1**: Counting mothers and babies per PHS-specified demographic criteria reveals the inherent diversity of our data set, given the obstetric population at our large, urban academic medical center.

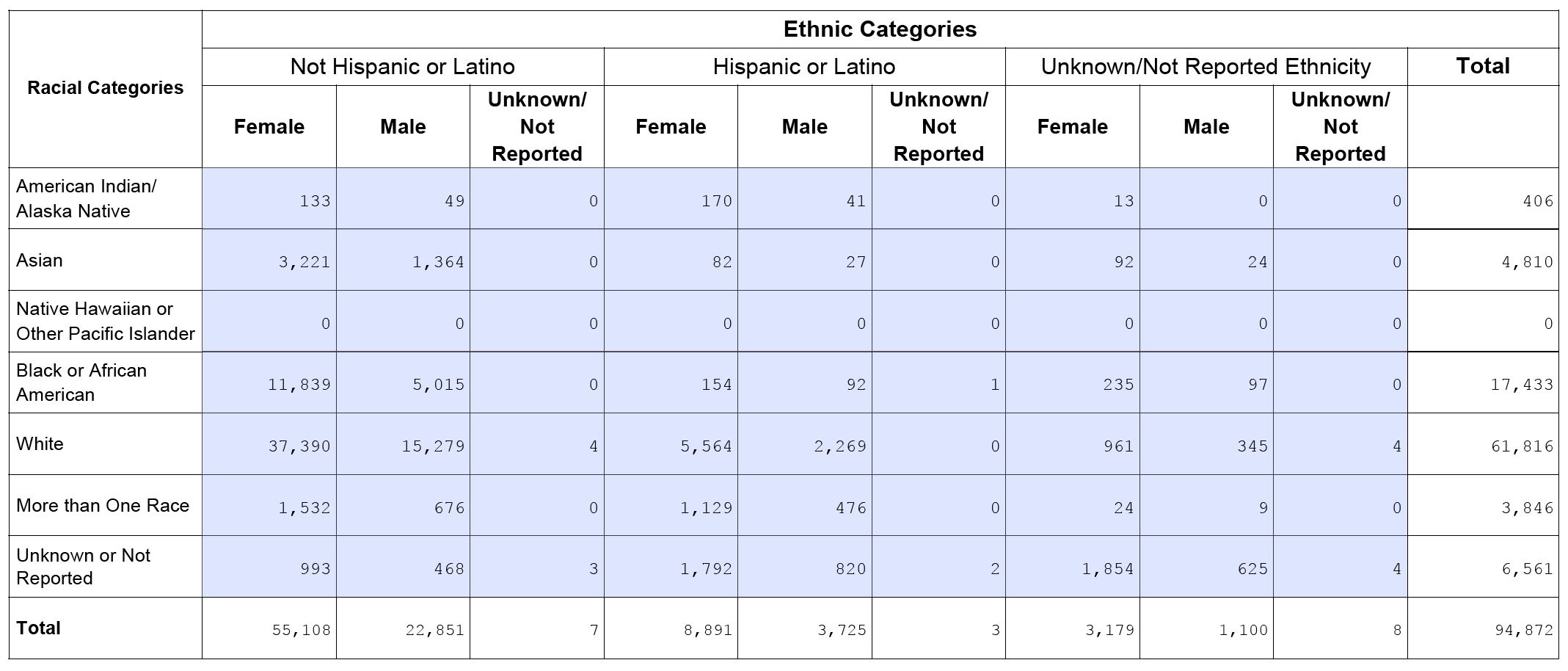

**References**

1.	The value of SNOMED CT. *SNOMED* https://www.snomed.org/snomed-ct/why-snomed-ct.

2.	OMOP Common Data Model – OHDSI. https://www.ohdsi.org/data-standardization/the-common-data-model/.

3.	Instructions and Form Files for PHS 398. https://grants.nih.gov/grants/funding/phs398/phs398.html.
